# Controlling the first wave of the COVID–19 pandemic in Malawi: results from a panel study

**DOI:** 10.1101/2021.02.21.21251597

**Authors:** Jethro Banda, Albert N. Dube, Sarah Brumfield, Amelia C. Crampin, Georges Reniers, Abena S. Amoah, Stéphane Helleringer

## Abstract

Many African countries have experienced a first wave of the COVID–19 pandemic between June and August of 2020. According to case counts reported daily by epidemiological surveillance systems, infection rates remained low in most countries. This defied early models of the potential impact of COVID–19 on the continent, that projected large outbreaks and massive strain on health systems. Theories proposed to explain the apparently limited spread of the novel coronavirus in most African countries have emphasized 1) early actions by health authorities (e.g., border closures) and 2) biological or environmental determinants of the transmissibility of SARS-CoV-2 (e.g., warm weather, cross-immunity). In this paper, we explored additional factors that might contribute to the low recorded burden of COVID–19 in Malawi, a low-income country in Southeastern Africa. To do so, we used 4 rounds of panel data collected among a sample of adults during the first 6 months of the pandemic in the country. Our analyses of survey data on SARS-CoV-2 testing and COVID-related symptoms indicate that the size of the outbreak that occurred in June-August 2020 might be larger than recorded by surveillance systems that rely on RT-PCR testing. Our data also document the widespread adoption of physical distancing and mask use in response to the outbreak, whereas most measured patterns of social contacts remained stable during the course of the panel study. These findings will help better project, and respond to, future waves of the pandemic in Malawi and similar settings.

## INTRODUCTION

In December 2019, a novel coronavirus, the severe acute respiratory syndrome coronavirus 2 (SARS-CoV-2), emerged in Hubei province, in Central China (1). Since then, SARS-CoV-2 has been detected in 191 countries. By the end of 2020, over 1.7 million deaths had been attributed to COVID–19, i.e., the disease caused by SARS-CoV-2 (2). The World Health Organization (WHO) declared this novel coronavirus a public health emergency of international concern on 30 January 2020 and classified its spread as a global pandemic on 11 March 2020. By comparison, the transmission of other coronaviruses (e.g., SARS-CoV-1, MERS-CoV) has been much more limited (3,4).

SARS-CoV-2 has however not spread evenly throughout the world. Europe and the Americas (e.g., USA, Brazil, Mexico) constitute the most affected regions (5). In these settings, the recorded incidence of the novel coronavirus has often remained high since the first few months of 2020, causing significant excess mortality relative to previous years (6–9). Other large outbreaks have been documented, for example in south Asia (10) as well as in Iran (11). In contrast, the incidence of SARS-CoV-2 appears low in African countries, which have recorded fewer than 4% of the global cases and deaths in 2020, despite accounting for more than 15% of the world population (12).

This limited burden of COVID–19 on the African continent contrasts with expectations and projections formulated at the beginning of the pandemic. At that time, there were strong concerns that overcrowding in slum neighborhoods of large cities, limited access to water and sanitation, inadequate human resources for health, and the high-prevalence of multi-generational households might create a particularly fertile ground for the spread of SARS-CoV-2 (13,14). A model from the WHO regional office for Africa thus projected 37 million symptomatic cases in 2020, requiring high levels of hospitalization and life support throughout the African continent (15). Whereas some African countries have indeed experienced multiple waves of the pandemic and high levels of excess mortality (e.g., South Africa), most African countries have not reported the predicted levels of strain on healthcare systems, and associated increases in morbidity and mortality (16,17).

Several theories have been proposed to explain the lower-than-expected burden of COVID–19 in most African countries in 2020. Some theories have emphasized the limited links between the continent and other world regions where SARS-CoV-2 first spread. African countries might thus have experienced a delayed onset of SARS-CoV-2 outbreaks due to the low volume of air travel to the region (18). Urbanization is also more limited in African countries than in other parts of the world (19). Lower population densities in rural areas limit opportunities for spreading SARS-CoV-2 in large regions of African countries (20). Other theories have suggested that the transmissibility of the novel coronavirus might be lower on most of the African continent. This might be because of climate factors, since SARS-CoV-2 spreads less efficiently in warm weather (21). It might also be attributed to (partial) protection from infection with SARS-CoV-2 due to prior exposure to other coronaviruses (17,21), genetic differences in susceptibility to viruses (22), or the non-specific effects of some vaccines (e.g., BCG) frequently administered to local populations (23). Furthermore, youthful age pyramids in African countries leave a smaller proportion of the population at-risk of severe COVID–19 and death compared to populations with an older age structure (20,24,25).

In addition, rapid policy responses might have played a key role in limiting the spread of SARS-CoV-2 in African countries. For example, several African countries closed their borders when COVID–19 cases were growing rapidly in China and in Europe. This might have delayed and/or reduced the importation of SARS-CoV-2. Restrictions on schools and gatherings and/or lockdowns might have reduced the scale on which the novel coronavirus could spread during the early phase of the pandemic. In South Africa, a lockdown and its associated travel restrictions might have limited the growth of the epidemic between March and June 2020 (26). Recent experience with other epidemic diseases (e.g., Ebola, HIV) might have better prepared local health actors for engaging local communities, and implementing required information campaigns, quarantines and contact tracing (27,28).

Beyond these oft-discussed factors, the limited burden of COVID–19 on the African continent might also be an artifact of incomplete data collection systems (12). As in other regions of the world, the recording of cases by epidemiological surveillance systems in African countries depends on the extent of testing for SARS-CoV-2 through RT-PCR. In several countries, the laboratory capacity to conduct such tests was not immediately available at the start of the outbreak and had to be established. In other countries, few reference laboratories had the capacity to carry out RT-PCR tests. These laboratories might have been initially overwhelmed by the volume of tests to be conducted to support pandemic response. Subsequently, the testing capacity has been greatly expanded in several countries, for example by mobilizing GeneXpert machines commonly used to test for tuberculosis (29). Nonetheless, African countries have so far conducted fewer tests per 1,000 people than elsewhere (30). The proportion of COVID–19 cases (and deaths) that are not recorded in case-based surveillance systems might thus be larger in African countries than in other parts of the world (12). Most African countries also lack death registration systems that are sufficiently complete to allow detecting peaks of excess mortality due to the pandemic, even in the absence of extensive testing for SARS-CoV-2 (31).

Even though governments were often quick to adopt recommendations from the WHO and the African CDC for the prevention of SARS-CoV-2 spread in local communities (e.g., physical distancing, enhanced hand hygiene, use of facial masks), few studies have investigated the contributions of such behavioral changes to epidemic control in African countries. Surveys conducted early in the pandemic in African countries, e.g., in the first few weeks after the introduction of SARS-CoV-2 in a country, have assessed the level of knowledge, and the perceptions of risk, about COVID-19 among various population groups (32–36). They have highlighted widespread concerns about getting infected with SARS-CoV-2, gaps in understanding of the modes of transmission of SARS-CoV-2, as well as varying levels of early adoption of protective behaviors (e.g., mask use, physical distancing). Subsequent surveys have focused predominantly on the impact of the COVID-19 pandemic on several socioeconomic dimensions including, for example, poverty, education, healthcare utilization, mental health and family planning (37–40).

In this paper, we analyzed data from a panel study conducted in Malawi during the 6 months that followed the introduction of SARS-CoV-2 in the country. We used these data to 1) evaluate the potential extent of under-reporting of COVID–19 cases in case-based surveillance systems, and 2) explore the pace at which protective behaviors (e.g., physical distancing, mask use) were adopted in response to the spread of SARS-CoV-2.

### BEHAVIORAL CHANGE TO CONTROL THE SPREAD OF SARS-COV-2

The potential for a pathogen like SARS-CoV-2 to spread within a population depends on the interactions between features of the virus and the characteristics and behaviors of its (human) hosts. This is summarized in an indicator called the basic reproduction number, *R*_0_ (41). It is defined as the number of new infections produced by a single infection in a completely susceptible population. An epidemic has the potential to expand when *R*_0_ is greater than 1, and it contracts when *R*_0_ is below 1. *R*_0_ is often expressed as the product of 3 parameters: (1) the probability that the virus is transmitted when a susceptible individual comes into contact with someone who is infected (transmissibility, noted *β*); (2) the average rate at which susceptible and infected individuals come into contact (noted *c̄*), and (3) the amount of time infected individuals remain contagious, i.e., they are able to transmit the virus (noted *d*). We thus have:

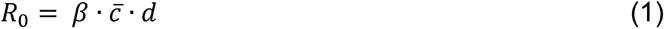

Controlling a viral outbreak then requires reducing one or several of those parameters, so that *R*_0_ drops below 1 in a sustained manner.

SARS-CoV-2 can be transmitted before the appearance, or in the absence, of symptoms (42–44). Because there are limited effective clinical options to reduce the duration of illness in mild or moderate COVID–19 cases (45), few control strategies target the parameter *d*. Instead, most recommended control strategies focus on altering *β* and/or *c̄*. Reducing the transmissibility of SARS-CoV-2 (*β*) can be achieved through vaccination, or through behavioral changes that limit exposure to viral particles emitted by infected individuals. This includes, for example, increasing hand washing to reduce transmission of SARS-CoV-2 through fomites (46,47), sufficient physical distancing to ensure that susceptible individuals cannot be reached by large infectious droplets (48), and wearing facial masks to reduce the risk of infection from droplets and other aerosols (49). Other interventions to reduce the transmissibility of SARS-CoV-2 may emphasize increasing ventilation of indoor space to limit transmission via aerosols (50).

Reducing the rate of contact between susceptible and individuals (*c*), on the other hand, requires encouraging or mandating that individuals limit their social interactions. Often, these injunctions target infected or exposed individuals: after a positive COVID–19 test, a presumptive diagnosis or a potential exposure to SARS-CoV-2, patients are asked to isolate so that they do not transmit the virus to others in their household, community or networks (51). In some circumstances, health authorities may also take wider measures to reduce the extent of contact within a population. They may temporarily prohibit attendance of certain places or events where individuals socialize and thus possibly transmit SARS-CoV-2. Since the beginning of the pandemic, many countries have even imposed periods of “lockdown” or “stay-at-home” orders, during which individuals are only allowed to leave their homes for restricted reasons and at very limited times.

In African countries, as in other areas of the world, governments and health stakeholders have promoted a mix of behavioral changes, which aim to reduce *β* and *c̄* simultaneously. Investigations of the determinants of epidemic dynamics in African countries have however focused on changes in the rate of contact between infected and susceptible individuals (*c̄*). Several mathematical models have projected the effects of lockdown measures on epidemic trajectories (52,53). Two empirical studies have relied on time-use surveys to document changes in patterns of contact in African settings (54,55). They used these data to build detailed age-specific contact matrices. In comparison with pre-pandemic data, they have shown that lockdowns reduced the extent of social interactions in target populations and prompted a decline in estimates of the reproduction number of the epidemic. Other mathematical models have explored the potential impact on epidemic trajectories in African countries of i) self-isolation of suspected COVID–19 cases, ii) contact tracing, and iii) “shielding” members of the most at-risk age groups (i.e., elderly people) (20,53,56). Several surveys have documented the prevalence of protective behaviors targeting *β*, i.e., the transmissibility of SARS-CoV-2, in African countries (32,33,54,57–59). However, they have seldom investigated whether the scale on which these behaviors are implemented might have changed over time.

### COVID–19 IN MALAWI

Malawi is a low-income country located in southeastern Africa, with a population of approximately 18 million. It had an estimated life expectancy of 63.7 years in 2019 (60). Since the 1980’s, it is affected by a large HIV epidemic (61,62). In 2015-16, more than 1 in 10 adults had HIV, and among those only 76.8% were aware of their HIV status (63). Prior to the COVID-19 pandemic, Malawi had made significant progress towards the achievement of various health-related goals, particularly those related to HIV treatment (64,65) and to the improvement childhood survival (66). In recent years, the incidence of non-communicable diseases linked to unhealthy behaviors has increased in urban and rural areas (67–69).

Initial projections of the potential impact of the COVID–19 pandemic in Malawi suggested that 1 in 5 Malawians might become infected with SARS-CoV-2 in 2020, resulting in more than 60,000 hospitalizations and close to 2,000 deaths (15). Malawi was one of the last countries in the world to record a case of COVID–19. This occurred on April 2^nd^ 2020, when infection was confirmed by RT-PCR in a traveler recently returned to the country (Figure 1). Sporadic clusters of COVID–19 cases were then detected in urban centers for several weeks. In late May, additional importations of COVID–19 cases occurred among migrants returning primarily from South Africa, i.e., the African country with the largest documented outbreak. The recorded incidence of SARS-CoV-2 then increased sharply in Malawi in June and July, before starting to decline in August. The daily number of new cases remained very low until the last weeks of December, when a sharp rebound in incidence was recorded in the country (figure 1). As of the end of December 2020, Malawi had recorded 6,583 cases of COVID–19, resulting in 189 deaths. The districts with the most confirmed cases were those where the country’s main cities are located (i.e., Lilongwe, Blantyre).

**FIGURE 1:**
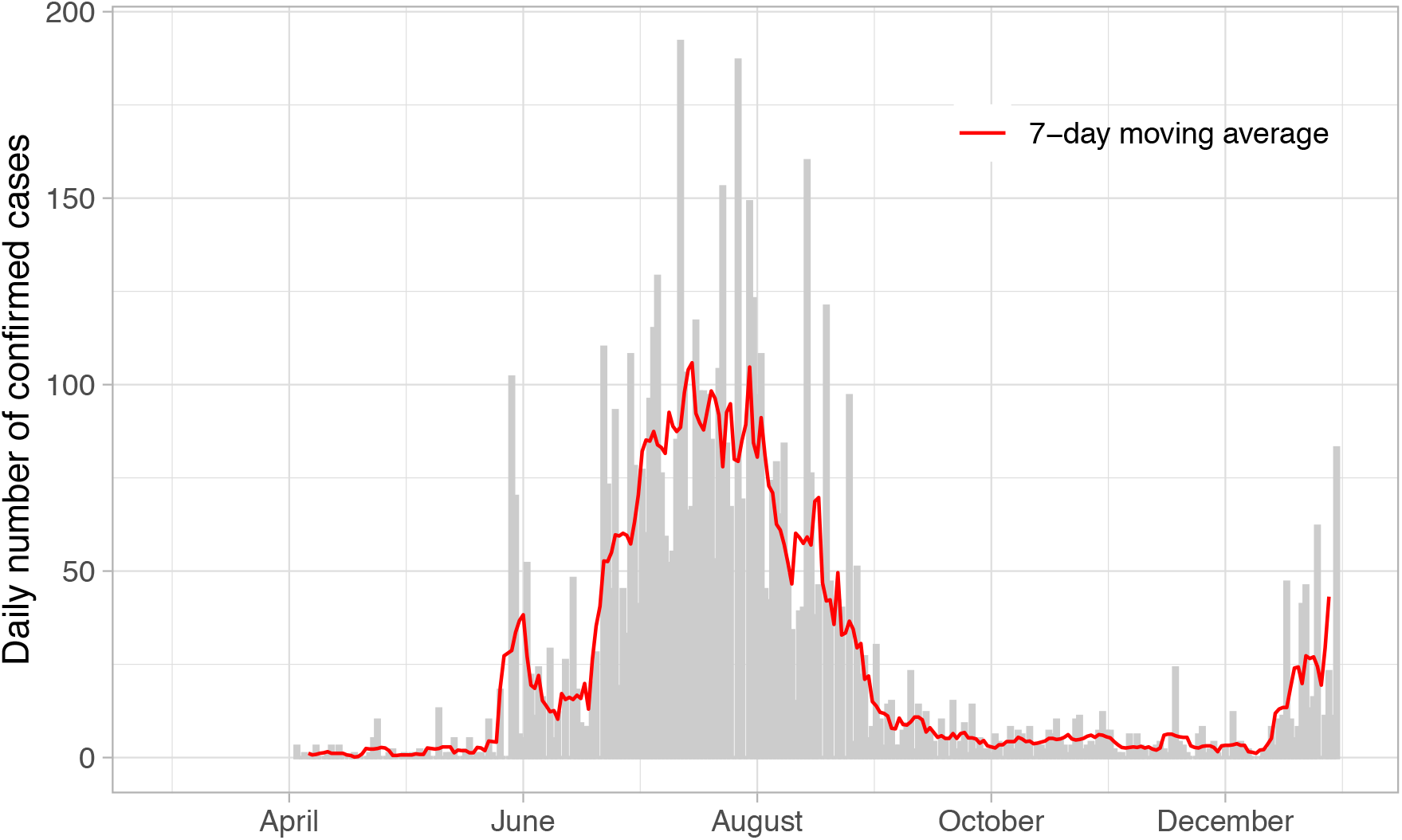
RECORDED CASES IN MALAWI. *Notes:* Data plotted in this graph were drawn from daily reports from the Public Health Institute of Malawi. The red line represents the 7-day moving average (calculated using the `zoo` package in R).

The Government of Malawi has adopted a series of measures to address the spread of SARS-CoV-2. At the end of March, prior to the emergence of the first COVID–19 cases in the country, the Government declared COVID-19 a “national disaster”, reduced entries into the country, and implemented COVID–19 screening at border posts and airports that remained open (e.g., to allow the entrance of essential goods). The Ministry of Health (MoH) and other stakeholders, initiated information campaigns (e.g., via radio messages, loudspeaker announcements or SMS) to increase awareness and knowledge of the pandemic among the population. This included communications about how SARS-CoV-2 spreads, information about possible symptoms, and what to do when presenting such symptoms. The MoH also started a toll-free hotline, as well as social media pages, where frequent updates about the epidemic situation in Malawi, and recommendations, were posted.

Laboratory-based surveillance systems similar to those used throughout the world were activated in the country (70). The capacity to detect SARS-CoV-2 by RT-PCR was established at the end of March at the National reference laboratory in Lilongwe, and at teaching and research institutions in Blantyre, in the southern region of the country (71). Additional laboratory sites with capacity to test for SARS-CoV-2 were then added throughout the country between May and July. As of the end of December 2020, Malawi had carried out more than 85,000 tests, i.e., slightly more than 4 tests per 1,000 people. In comparison, neighboring countries have conducted tests on a larger scale (e.g., 9.6 tests per 1,000 in Mozambique, 38.6 tests per 1,000 in Zambia), whereas in high-income countries, testing rates often exceed 200 tests per 1,000 (30).

COVID-19 tests have been carried out in various contexts in Malawi, including screening of travelers at border posts and diagnostic tests for suspected cases identified in health facilities. Tests have also been conducted as part of contact tracing (72), i.e., efforts through which health workers get in touch with people who have come in contact with a confirmed case of COVID–19, notify them of their exposure, and encourage them to isolate in an attempt to break transmission chains. Between the beginning of the outbreak and the end of 2020, more than 10,000 contacts of COVID–19 cases have been listed and followed-up in the country.

In Malawi, the MoH and other stakeholders (e.g., organizations of the UN system, Non-Governmental Organizations) have sought to reduce the transmissibility of the novel coronavirus using various channels. Protective behaviors such as increased hand washing, coughing or sneezing in one’s elbow, and physical distancing were thus rapidly promoted. Early in the outbreak, the MoH also emphasized the use of facial masks to prevent the transmission of SARS-CoV-2. Medical masks and respirators were recommended in healthcare settings, and among people who care for patients with COVID-19, whereas the general public was advised to use cloth masks “in settings where social distancing is not possible and where there is widespread community transmission” (73). The recommendations were accompanied by guidelines intended for local tailors about the making of cloth masks (e.g., number of fabric layers). To respond to the increased spread of SARS-CoV-2 in June and July (see figure 1), the Government made mask wearing mandatory when in public in early August. The measure was accompanied by a fine of 10,000 Malawian Kwachas (i.e., approximately 12 USD) for non-compliers.

Concurrently, several measures were taken to reduce contact between susceptible and infected individuals in the population. As part of the national disaster declaration at the end of March, schools and universities were closed throughout the country, and attendance of public gatherings was limited to 100 people. A national lockdown was then announced on April 18^th^ (74). However, several organizations contested the proposed plan. Following legal procedures, the Malawi High Court suspended the implementation of this “stay-at-home” order. Unlike other countries in Eastern and Southern Africa (26), Malawi thus never experienced a nationwide lockdown in 2020. The government of Malawi nonetheless adopted other measures to reduce contacts in specific populations. For example, some prisoners were released to help reduce overcrowding, and thus prevent large outbreaks, in prisons in August 2020. In recent months, some of the restrictions on social contact have been lifted in Malawi. For example, schools and universities reopened in September. International travel (e.g., flights) also resumed on a larger scale at that time, before being interrupted again in December in response to increased incidence of SARS-CoV-2 (figure 1).

## DATA AND METHODS

### Panel dataset

To evaluate potential explanations for the low observed burden of COVID–19 in Malawi, we analyzed panel data collected during the first 6 months of the pandemic in the country (“COVID–19 panel” thereafter). This panel was initiated in April 2020, roughly a month after the Government of Malawi declared COVID–19 a “national disaster”, and less than 3 weeks after the first cases of COVID–19 detected in Lilongwe, the capital city. The COVID–19 panel now includes 4 rounds of data collection conducted approximately 6 weeks apart and ending in mid-November 2020. During that time, the recorded incidence of the SARS-CoV-2 virus varied greatly, with peaks of close to 200 cases per day in July vs. fewer than 5 cases per day on average in October and November (figure 1). The first two rounds of data collection also took place during a time when campaigning for the presidential election of June 23^rd^ was under way (75).

Respondents in the COVID–19 panel had previously participated in a study of the measurement of adult mortality. This “pre-COVID–19 study” was completed before the introduction of SARS-CoV-2 in Malawi. It was nested within the activities of a health and demographic surveillance system (HDSS) located in Karonga district, in northern Malawi (76,77). This is an area bordering Lake Malawi to the east and located within 1.5-hour drive of the Tanzania border. Its economy is predominantly organized around fishing, subsistence farming (e.g., rice, cassava) and small-scale retail activities.

The last round of interviews for the pre-COVID–19 study focused on the health and family relations of respondents. It occurred between December 2019 and March 2020. At that time, we sought to obtain the mobile phone numbers of 1) a random sample of current residents of several HDSS population clusters located close to the lakeshore community of Chilumba (“current residents”, thereafter), 2) a random sample of former HDSS residents who had migrated throughout Malawi (“former residents”, thereafter), and 3) the siblings of these current and former HDSS residents (“referred siblings”, thereafter). When the spread of SARS-CoV-2 accelerated throughout the world (including in other parts of Africa) in March 2020, we decided to initiate the COVID–19 panel: we contacted all participants of the pre-COVID-19 study for whom a phone number was available, and invited them to participate in a series of COVID-related interviews.

### Study sample

In total, we sought to obtain the phone numbers of 1,036 participants in the pre-COVID–19 study and were successful for 779 participants (75.2%). Among those approximately 80% agreed to enroll in the COVID–19 panel (n = 619). More detailed descriptions of the constitution of the COVID–19 panel are available elsewhere (78). Compared to other participants in the pre-COVID–19 study, those who joined the COVID–19 panel were more educated (78). Follow-up rates from one round of the COVID–19 panel to the next were 93-95%, and in total, 543 respondents completed all 4 rounds of the COVID–19 panel (figure 2).

**FIGURE 2:**
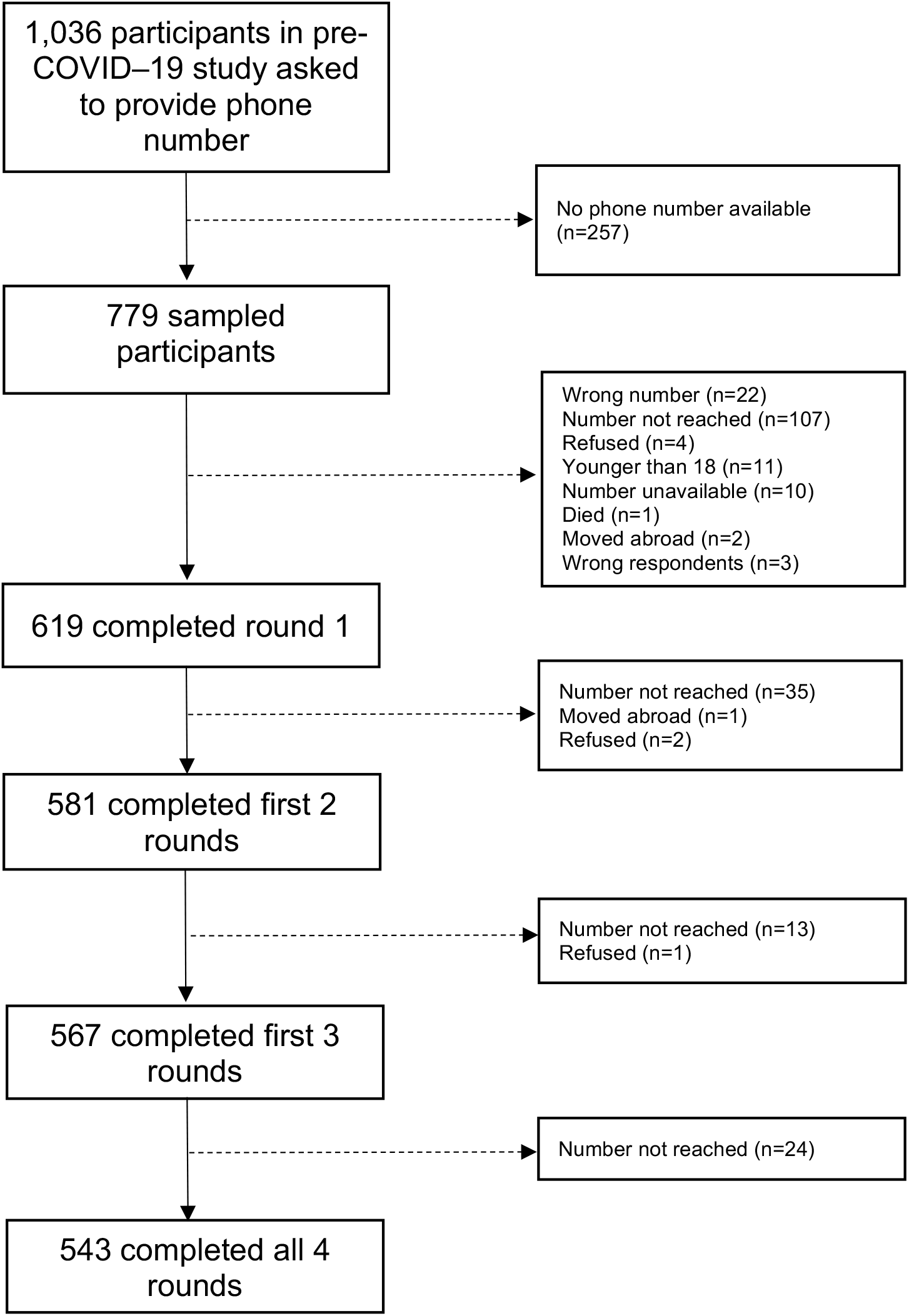
FLOW CHART OF STUDY PARTICIPATION.

Participants in the COVID–19 panel included men and women aged 18 years and older. The majority of respondents were residents of rural and peri-urban parts of Karonga district (see appendix A1). Other participants in the COVID–19 panel were dispersed throughout the country (appendix A1), including in districts and large urban centers of the central and southern regions.

Due to its sampling frame, and to selective participation in a survey conducted by mobile phone, the sample of the COVID–19 panel is not representative of the population of Malawi, or Karonga district. We thus do not use these data to estimate the prevalence of various symptoms or behaviors in the country at various points during the pandemic. Instead, we use multiple measurements available for each panel participant to a) understand the potential scale of under-reporting of COVID–19 in data derived from RT-PCR tests, and b) assess the pace at which study participants adopted protective behaviors against SARS-CoV-2.

### Data collection

All interviews of the COVID–19 panel were conducted by mobile phone, by a team of trained interviewers. This team had previously participated in data collection of the most recent Malawi Demographic and Health Survey (2016), as well as in the pre-COVID–19 study. Each interviewer was randomly assigned a set of respondents at the beginning of the COVID–19 panel, and these sets were maintained in all 4 rounds of data collection. As in other surveys conducted during the COVID–19 pandemic in African countries (e.g., 79), data collectors conducted interviews from their own homes. We sought oral informed consent from all participants before each round of data collection. Respondents who completed an interview were provided with 1,200 Malawian Kwachas worth of mobile phone units (approximately 1.6 US Dollars). Data collection procedures for the COVID–19 panel were approved by the institutional review boards of the National Health Sciences Research Committee in Malawi, the Johns Hopkins University School of Public Health and the London School of Hygiene and Tropical Medicine.

### Measuring COVID-related symptoms and behaviors

In each round of the COVID–19 panel, we asked respondents whether they had experienced, during the previous month, a number of symptoms previously reported among COVID–19 cases in clinical studies (11,80,81). These symptoms included, coughing, shortness of breath, chest pain, fever, headaches or fatigue. We only included loss of smell/taste (82,83) in the list of symptoms elicited in rounds 2, 3 and 4 of the COVID–19 panel. Starting in round 3, we asked respondents who reported any symptom possibly related to COVID–19 whether they self-isolated upon experiencing these symptoms. We also asked respondents whether they had ever been tested for SARS-CoV-2. In round 4, we added a description of the sample collection procedure in our question about SARS-CoV-2 testing (i.e., that a long swab was inserted in the patient’s nostrils). This description was introduced to help prevent confusion with other COVID-related screening procedures (e.g., temperature checks) widely in Malawi implemented since the start of the pandemic.

In each round of data collection, we investigated the behaviors respondents used to reduce the transmissibility of SARS-CoV-2 and/or limit their contacts in the month before the survey. First, to explore changes in *c*, i.e., the rate of contact between infected and susceptible individuals, we inquired about respondents’ living arrangements. We asked respondents how many people resided in their households, and how many rooms their house included. This provided an indicator of the potential for intra-household exposure to/transmission of SARS-CoV-2. Second, to measure contacts outside of the household, we asked respondents whether they had attended various places and events. We drew a list of places where people commonly interact and socialize in Malawi, which included markets, churches and mosques, neighbors or relatives’ houses, workplaces, football games or funerals. Then, we asked respondents if they had attended each of these places/events in the past 7 days before the interview.

We also asked respondents to list the behaviors they had used to reduce the spread of SARS-CoV-2 in the past month. This question was adapted from instruments previously administered in COVID-related surveys conducted in high-income countries (84). During pre-testing and piloting activities, we drew a list of possible answers to this question. This list included behaviors that reduce *β*, e.g., use of facial masks, physical distancing or handwashing, as well as behaviors that reduce *c̄*, e.g., avoiding crowded areas. Interviewers did not read this list to respondents. Instead, they let them spontaneously recall their behaviors, and coded their answer using the list of potential behaviors. If a behavior was not included in the list, they coded it as “other” and specified the respondent’s answer in a follow-up question. Multiple answers were allowed, and after each answer, interviewers were instructed to probe further in a non-specific manner, by asking respondents whether there was anything else that they did to prevent the spread of SARS-CoV-2.

In rounds 3 and 4, we asked respondents who reported wearing a facial mask in the past month how often they wore one during that timeframe. Possible answers were “always”, “often”, “sometimes” and “rarely”. We also asked them to list the reasons why they wore a mask. Interviewers let respondents answer spontaneously and they coded their answer using a list of possible reasons. This list included, for example, “self-protection”, “to protect others”, “to meet a requirement (e.g., to enter a building)” or “to avoid a fine”. Multiple answers were allowed. After each reason mentioned by a respondent, interviewers were instructed to probe further by asking whether there was an additional reason.

### Data analysis

we first described the socio-economic characteristics of panel participants who completed all 4 rounds of data collection, including their reported gender, age, marital status, occupation and religion. Second, we analyzed trends in their experience of COVID-related symptoms over time. We classified participants in 3 groups according to their reported experience of symptoms during the course of the study: those without any reported symptoms, those who developed a cough, and those who developed one or more other symptoms previously associated with COVID-19. We then assessed the level of testing for SARS-CoV-2 in our panel sample, and we explored whether testing rates varied across these 3 groups. To measure testing rates, we only considered SARS-CoV-2 testing histories reported during round 4, because we introduced a more precisely worded question during that round. Low testing rates among those with new onset of various COVID-related symptoms constitute an indication of the extent of under-reporting of the SARS-CoV-2 burden in this sample.

Third, we described trends in behaviors associated with social contacts in our panel sample. To assess the potential for intra-household contact, we reported the ratio of the number of household residents to the number of rooms in the house. Then, we tabulated reports of attendance of places and events in the past week collected during each round. We measured the proportion of respondents reporting that they self-isolated when they experienced symptoms indicative of COVID–19, and we tested whether the likelihood of self-isolation varied depending on the type of symptom experienced. We tabulated other reported behaviors, which might reduce the rate of contact between infected and susceptible individuals, by data collection round.

Finally, we measured trends in reported behaviors related to the transmissibility of the virus (*β*) across all 4 rounds. We included all such behaviors listed in response to our open-ended question about behaviors used to prevent SARS-CoV-2 spread. For respondents who reported using facial masks in rounds 3 & 4, we also measured the reported frequency of mask use, and we described the reasons they listed for using facial masks.

Data compiled by the MoH suggest that SARS-CoV-2 spread more extensively in large cities in Malawi. We thus conducted all analyses of the panel dataset separately for residents residing in urban vs. rural areas. We used the definition of “urban” areas from the Malawi National Statistical Office: “urban centers in Malawi refer to the four major cities [of] Blantyre, Lilongwe, Mzuzu, and Zomba, town councils and bomas^6^ and other town-planning areas” (85). All other areas of residence were classified as “rural”. All analyses were conducted using the statistical software, R.

## RESULTS

### Descriptive statistics

There were 543 participants who completed the four rounds of the COVID–19 panel (figure 2). Among those, 419 resided in rural areas at round 1, whereas 124 resided in urban areas (table 1). In this sample, there were few respondents aged 55 and older (approximately 2%), whereas close to 1 in 5 full participants was aged 18–24 years old. There were large differences in the marital status, water source, occupation and religion of residents residing in rural vs. urban areas. Rural participants were more likely to be currently married (64.0%) or formerly married (21.9%) than urban participants (54.8% and 10.4%, respectively). Rural participants relied more frequently on boreholes as their primary water source (55.1% vs. 4.8% in urban areas) and they engaged in activities related to agriculture and fishing at a much higher rate (46.0% vs. 8.6% in urban areas).

**TABLE 1:** CHARACTERISTICS OF PANEL PARTICIPANTS REPORTED DURING ROUND 1 OF DATA COLLECTION, URBAN VS RURAL. *Notes:* all figures in parentheses are column percentages; ^1^ Occupation was measured during round 2 of the COVID–19 panel. ^2^ Religion was measured during round 3 of the COVID–19 panel. ^3^ CCAP = Church of Central Africa Presbyterian.

| Area of Residence | Rural | Urban |
| --- | --- | --- |
| Sex of the respondent |  |  |
| Male | 169 (40.3%) | 48 (38.7%) |
| Female | 250 (59.7%) | 76 (61.3%) |
| Age |  |  |
| 18-24 | 76 (18.1%) | 25 (20.2%) |
| 25-34 | 144 (34.4%) | 42 (33.9%) |
| 35-44 | 133 (31.7%) | 41 (33.1%) |
| 45-54 | 55 (13.1%) | 14 (11.3%) |
| ≥55 | 11 (2.6%) | 2 (1.6%) |
| Marital status |  |  |
| Currently married | 268 (64.0%) | 68 (54.8%) |
| Separated | 55 (13.1%) | 5 (4.0%) |
| Divorced | 21 (5.0%) | 4 (3.2%) |
| Widowed | 16 (3.8%) | 4 (3.2%) |
| Never married | 59 (14.1%) | 43 (34.7%) |
| Water source |  |  |
| Piped water in house | 74 (17.7%) | 69 (55.6%) |
| Piped water - shared | 65 (15.5%) | 44 (35.5%) |
| Borehole | 231 (55.1%) | 6 (4.8%) |
| Lake/River | 49 (11.7%) | 5 (4.0%) |
| Occupation<sup>1</sup> |  |  |
| Professional/technical/managerial | 15 (3.6%) | 29 (22.7%) |
| Sales and services | 102 (24.6%) | 37 (28.9%) |
| Manual work | 57 (13.8%) | 24 (18.8%) |
| Domestic service | 50 (12.0%) | 27 (21.1%) |
| Agriculture/fishing | 191 (46.0%) | 11 (8.6%) |
| Religion<sup>2</sup> |  |  |
| Catholic | 73 (17.5%) | 26 (20.8%) |
| CCAP<sup>3</sup> | 81 (19.4%) | 41 (32.8%) |
| Pentecostal | 83 (19.9%) | 28 (22.4%) |
| Other Traditional churches | 140 (33.5%) | 15 (12.0%) |
| Other | 41 (9.8%) | 15 (12.0%) |

### Symptoms and SARS-CoV-2 testing

The proportion of respondents reporting various symptoms associated with COVID–19 varied during the course of the panel study. The proportion of rural participants reporting a cough in the month prior to the survey thus declined steadily from 28.2% in round 1 to 18.1% in round 4 (figure 3). Among urban participants, on the other hand, the proportion of respondents reporting a cough in the previous almost doubled between rounds 1 & 2 (from 17.7% to 30.5%), before returning to lower levels in rounds 3 and 4 (16.0% and 17.8%, respectively). Other symptoms associated with COVID–19 displayed similar trends among respondents in urban areas. For example, no urban participants reported chest pain in round 1 of the panel vs 5% in round 2, and <2% in rounds 3 and 4 (not shown).

**FIGURE 3:**
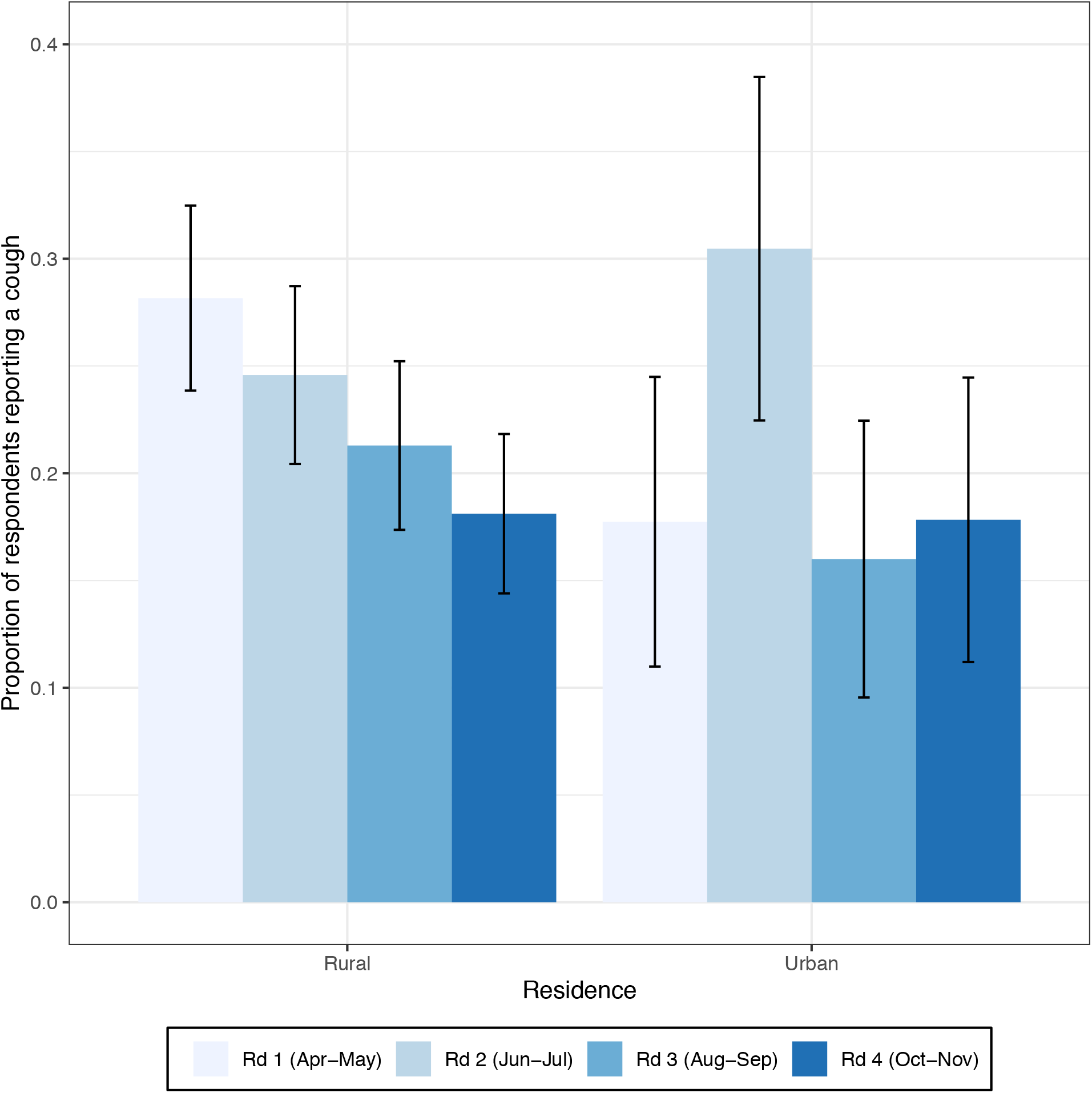
PREVALENCE OF REPORTED COUGH, URBAN VS RURAL. *Notes:* the survey question asked respondents whether they experienced a cough in the past month prior to the survey. Estimates of the standard errors (used to calculate confidence intervals) were adjusted for the clustering of observations within families.

Few participants (5.9%) reported being tested for SARS-CoV-2 since the start of the pandemic in Malawi. Among rural participants, the likelihood of SARS-CoV-2 testing was not associated with experience of symptoms related to COVID–19 (figure 4), whereas among urban participants, the likelihood of SARS-CoV-2 testing was slightly higher among those who had developed a cough or other symptoms associated with COVID–19 during the course of the study. Rural participants reported being tested at health facilities or at home (as part of contact tracing efforts), whereas a few urban participants also reported being tested at border posts.

**FIGURE 4:**
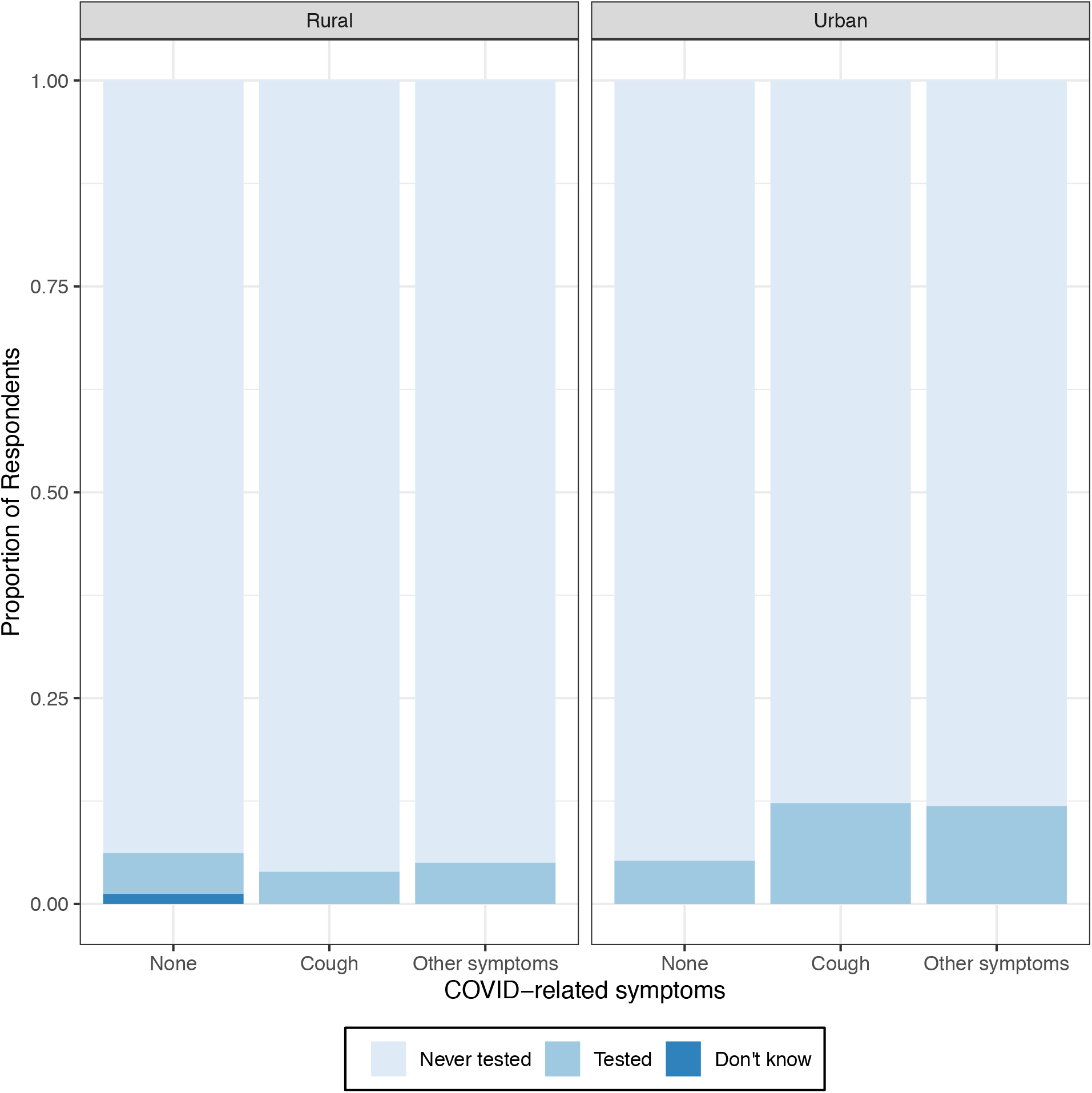
COVID-19 TESTING ACCORDING TO EXPERIENCE OF COVID-RELATED SYMPTOMS. *Notes:* The experience of symptoms was ascertained over the course of the panel study, whereas the experience of testing for SARS-CoV-2 was reported in round 4. COVID-related symptoms elicited during the surveys included fever, chest pains, shortness of breath, fatigue, shivering, headache, muscle pain, runny nose, sore throat, cough and wheezing.

***Changes in contact patterns (*c̄*):*** there were limited changes in the living arrangements of panel participants during the course of the study (figure 5). Both rural and urban participants reported living in households with a median density of approximately 2 people per room. Crowded households (i.e., with more than 3 people per room) were slightly more common in rural areas. In addition, <1% of respondents reported using residential strategies (e.g., moving to a new house or flat, appendix A2) to reduce the spread of SARS-CoV-2.

**FIGURE 5:**
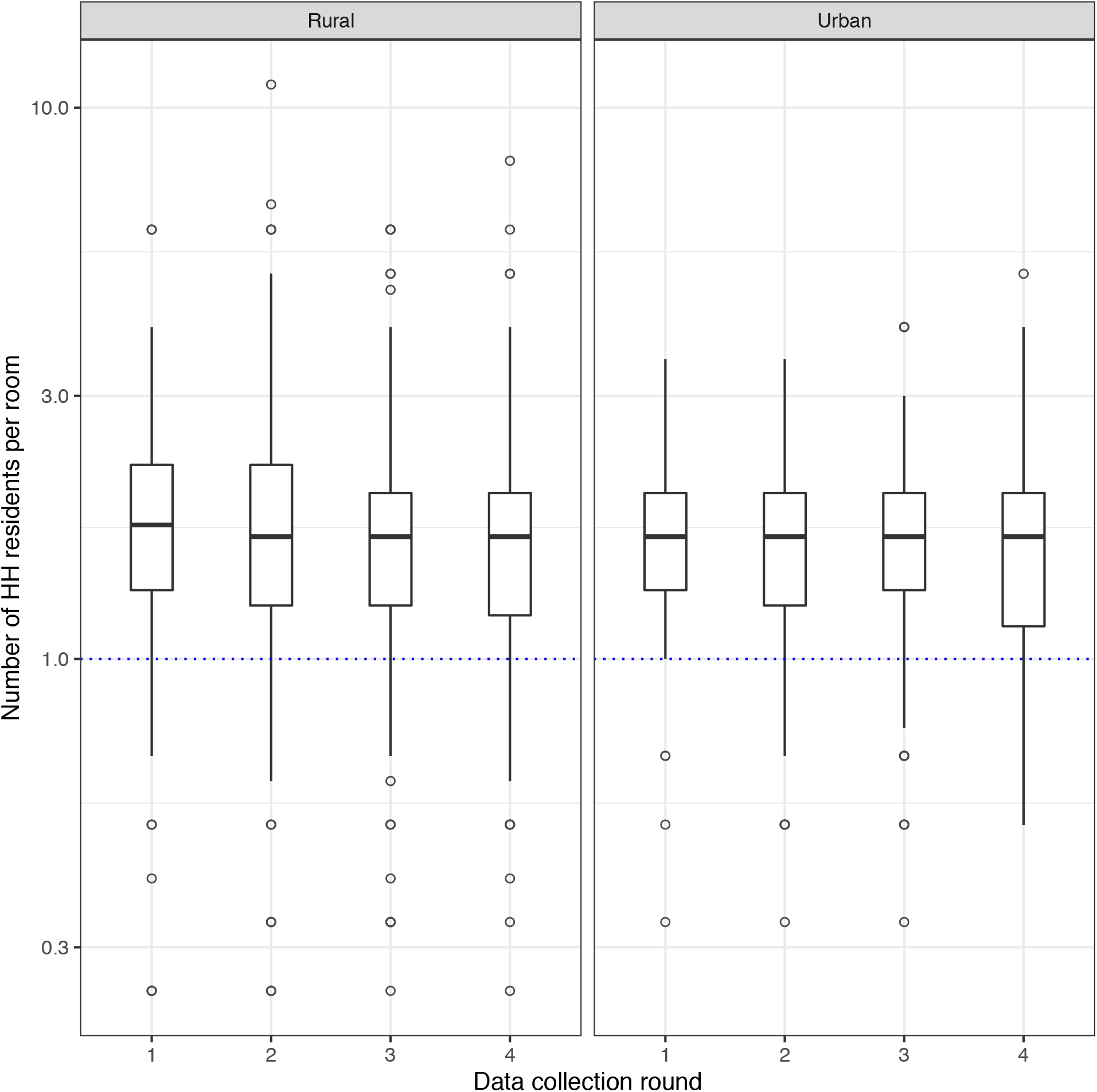
CHANGES IN HOUSEHOLD OCCUPANCY RATIO. *Notes:* the y-axis is plotted on a logarithmic scale. The results presented in this graph are derived from two survey questions asking respondents 1) how many people reside in their household, and 2) how many rooms there are in their house. In each round, the median number of residents per room was 2, both in rural and urban places.

There were limited changes in attendance of places and events where people socialize across all 4 rounds. Rural and urban participants reported most commonly visiting markets, their neighbors’ or relatives’ homes, places of worship and workplaces (figure 6). The proportion of rural participants reporting attending a funeral in the past 7 days declined slightly after round 1 of the panel, from 47.5% in round 1 to approximately 37% in rounds 2, 3 and 4. The proportion of rural participants attending football games varied between rounds, from 17.9% in round 1 to 38.3% in round 2. On the other hand, attendance of schools/universities increased sharply between rounds 3 and 4: among participants in rural areas, the proportion of participants visiting a school in the past 7 days increased from 3.6% in round 1 to 13.8% in round 4; among urban residents, similar figures were 3.2% and 27.9%. Very few participants reported attending weddings and baptisms in the early rounds of the panel. However, the proportion of urban residents who reported attending a wedding in the past week increased from <1% in round 1 to 17.8% in round 4.

**FIGURE 6:**
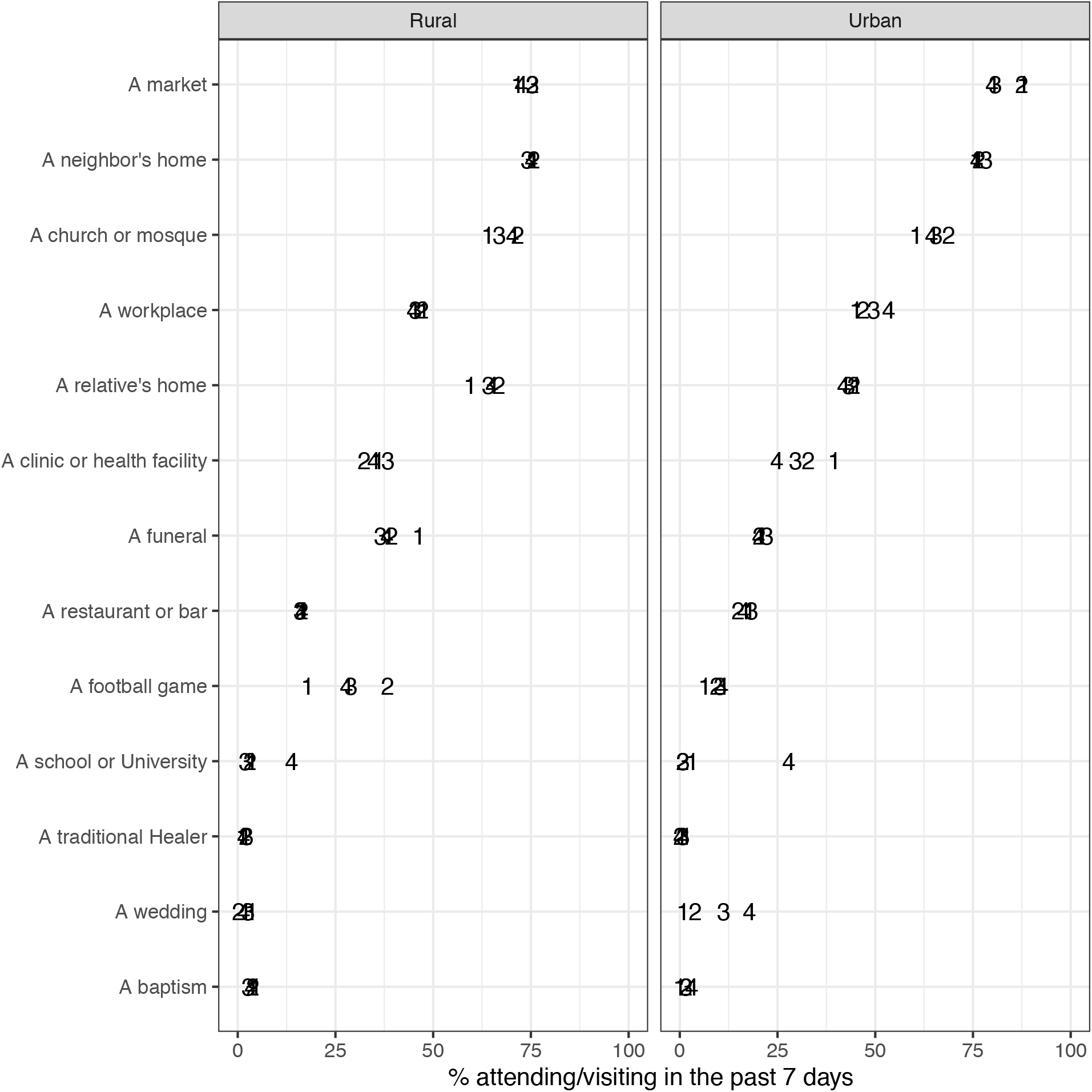
ATTENDANCE OF PLACES & EVENTS. *Notes:* the data plotted in this graph were collected from a series of questions asking respondents whether they had attended the places listed on the y-axis in the past 7 days prior to the survey. The point estimates are labeled by their round number on the plot. The places that appear on the y-axis are ordered according to their attendance in urban areas in round 1.

Compliance with isolation guidelines was limited among panel participants (figure 7). The proportion of participants experiencing COVID-related symptoms who reported self-isolating was 29.1% in round 3 vs. 24.5% in round 4. The likelihood of self-isolation also varied by a) place of residence, and b) the type of COVID-related symptom experienced. In particular, participants who experienced a cough were more likely to report self-isolating than participants who reported other symptoms (e.g., fever, headaches). Self-isolation was especially high during round 3 among urban participants who had recently been coughing (i.e., 70%).

**FIGURE 7:**
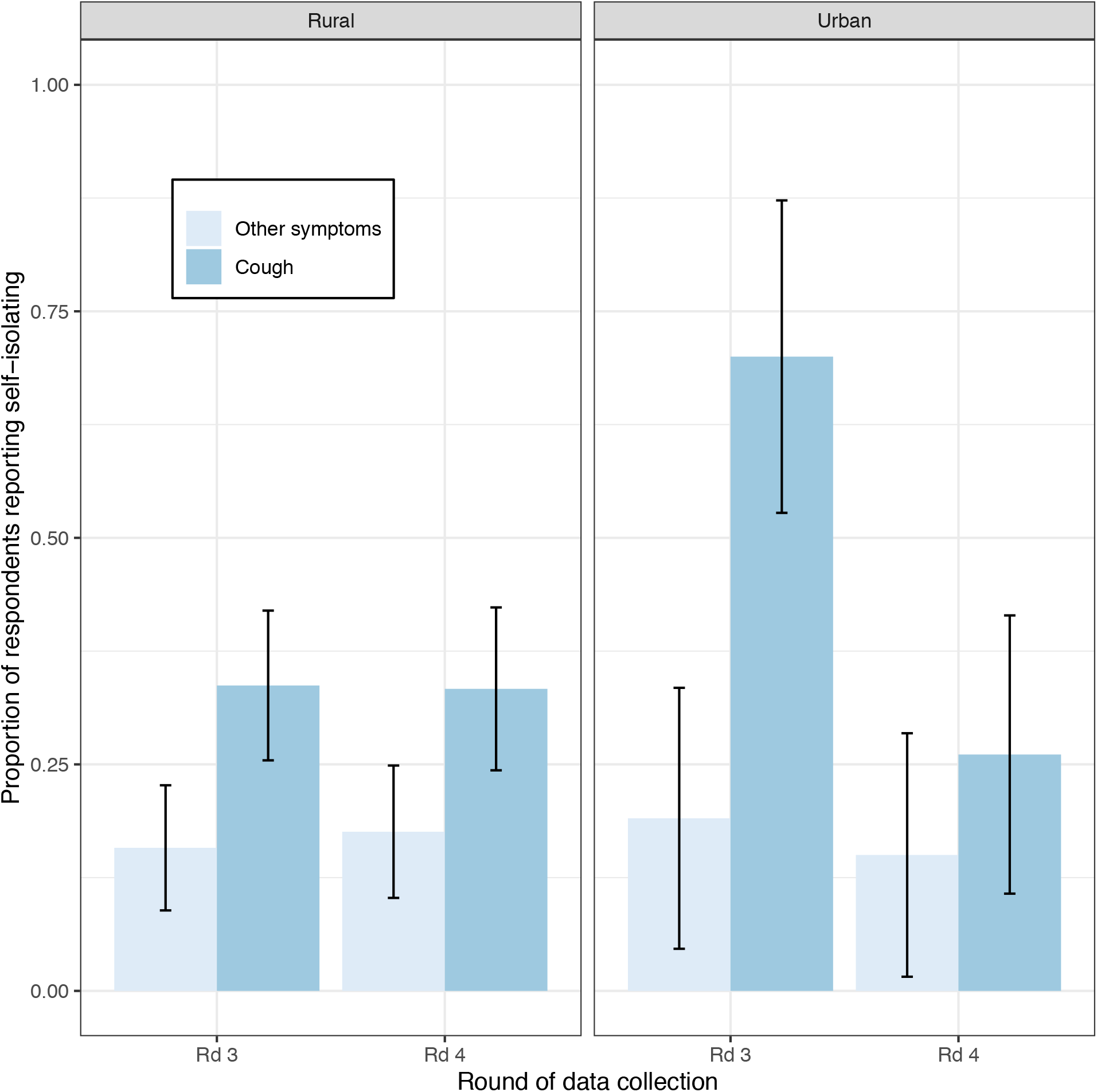
SELF-ISOLATION BEHAVIORS, BY EXPERIENCE OF SYMPTOMS. *Notes:* the question about self-isolation was only applicable for respondents who reported experiencing one or more COVID-related symptoms in the month prior to the survey. This question was introduced in round 3 of the panel survey. In round 3, the sample size for these analyses was n = 206. In round 4, the sample size was n = 192. Estimates of the standard errors (used to calculate confidence intervals) were adjusted for the clustering of observations within families.

Some panel participants also reported “avoiding crowded areas” or “avoiding going out in general” to reduce the spread of SARS-CoV-2 (appendix A2). However, the proportion of respondents using these strategies to reduce contact declined after round 1. Very few respondents also reported reducing their mobility to prevent the spread of SARS-CoV-2 (appendix A2). In round 1, >10% of panel participants in urban areas reported “avoiding visibly sick people” to prevent the spread of SARS-CoV-2, but in subsequent rounds, this proportion declined to <2%. Virtually none of the panel participants reported “avoiding healthcare facilities” to reduce their risk of acquiring or transmitting SARS-CoV-2.

***Changes in behaviors related to transmissibility (β):*** panel participants reported adopting various behaviors that modify the transmissibility of SARS-CoV-2. In each round, more than 90% of participants reported washing their hands more often (figure 8). In round 1, the next most commonly reported behaviors affecting transmissibility were the use of hand sanitizer among urban respondents and covering up coughs/sneezes among rural residents. In subsequent rounds however, panel participants reported adopting other strategies as their main preventive behaviors. For example, whereas the use of physical distancing was reported by approximately 10% of participants in round 1, it increased to close to 60% in round 2 among participants in urban areas, and to similar levels in round 3 among participants in rural areas. The use of physical distancing then stopped increasing in rural areas in round 4, and even declined among urban participants.

**FIGURE 8:**
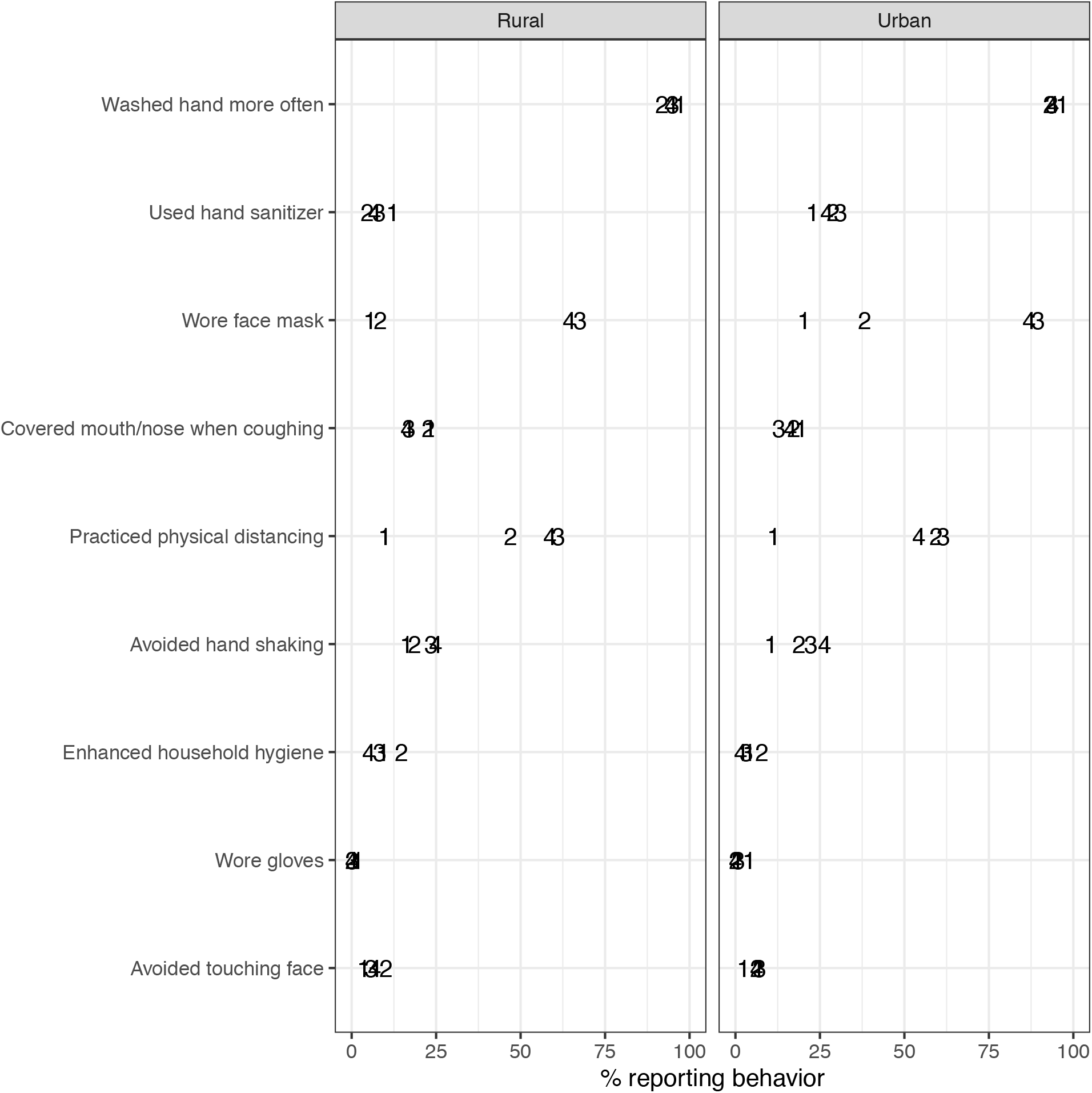
ADOPTION OF BEHAVIORS TO REDUCE TRANSMISSIBILITY OF SARS-COV-2. *Notes:* the data plotted in this graph were collected from a question asking respondents what they had done in the past month to prevent the spread of SARS-CoV-2. Respondents were not provided with potential answers. However, after each response, interviewers were instructed to probe further, by asking respondents: “is there anything else that you have done?”. In this graph, the behaviors listed on the y-axis are those that reduce the transmissibility of the coronavirus. Respondents also listed other behaviors, that reduce the contact between infected and susceptible individuals (appendix A3). In each round, <1% of panel respondents reported not doing anything to prevent the spread of SARS-CoV-2. The point estimates are labeled by their round number on the plot. The behaviors that appear on the y-axis are ordered according to their prevalence in urban areas in round 1.

In all rounds, the use of facial masks in the month prior to the survey was more widespread among participants in urban areas than in rural areas (figure 8). Reported mask use did not increase markedly between rounds 1 (4.4%) and 2 (7.1%) among participants in rural areas, whereas the proportion of urban participants who reported using a mask almost doubled during the same timeframe (20.3% in round 1 vs. 38.3% in round 2). The proportions of participants reporting use of facial masks in the month prior to the survey then increased particularly sharply in subsequent rounds: in round 3, 89.6% of participants in urban areas, and 67.7% in rural areas, reported using a mask in the past month.

The proportion of participants who reported using a mask at any point in the past month remained approximately constant in rounds 3 & 4 of the panel study. However, the frequency of mask use declined markedly between round 3 and 4 (figure 9). In rural areas, the proportions of mask users who reported wearing masks either “always” or “very often” declined from 71.2% to 39.7%, whereas in urban areas, corresponding figures were 78.9% and 44.7%.

**FIGURE 9:**
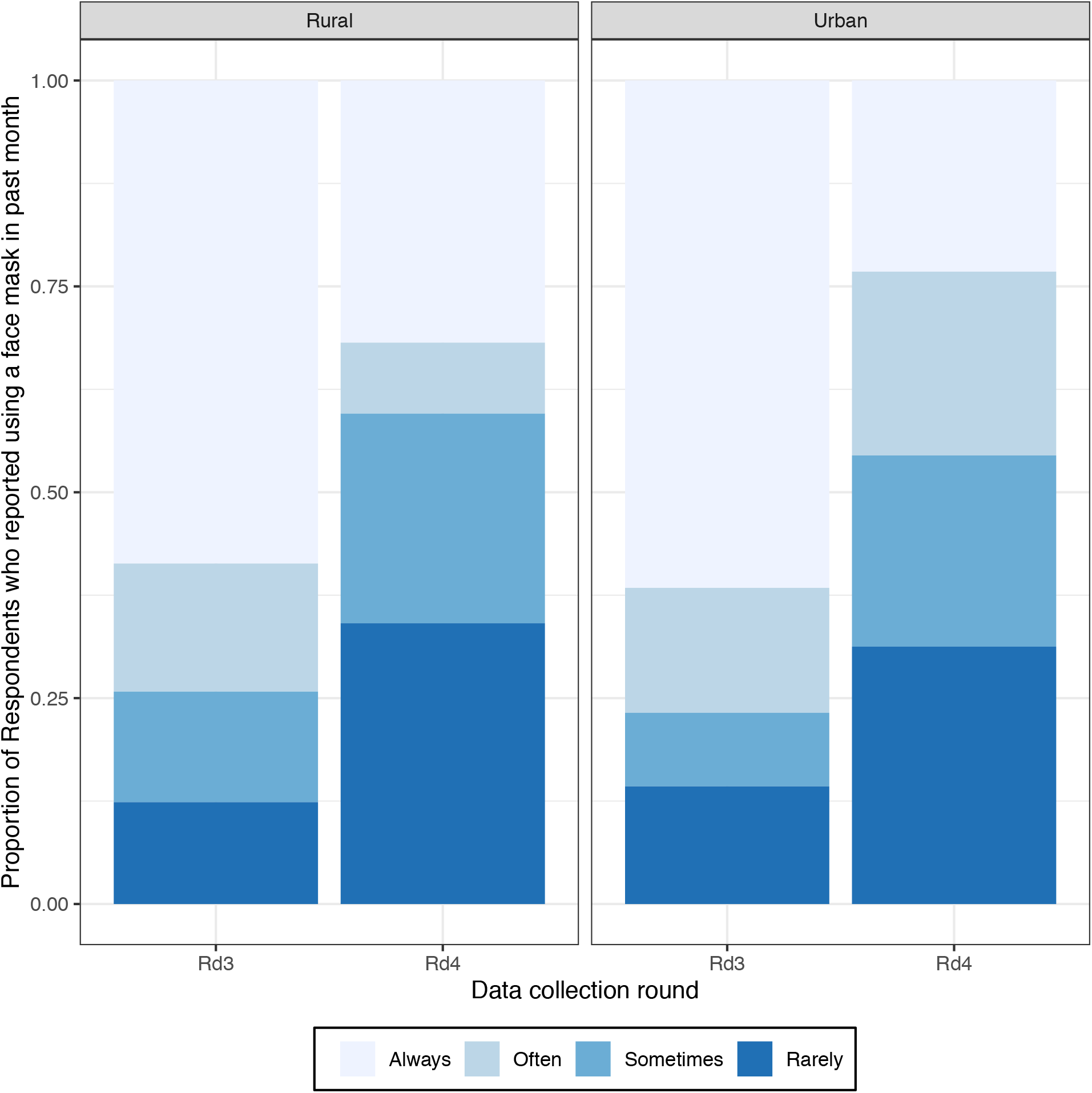
FREQUENCY OF MASK USAGE. *Notes:* in rounds 3 & 4, we asked respondents who reported that they had used a mask to prevent the spread of SARS-CoV-2, how often they had done so. These data were not collected in rounds 1 & 2. In round 3, the sample size for these analyses was n = 498. In round 4, the sample size was n = 496.

The reasons for wearing masks reported by participants also varied between rounds 3 and 4. In round 3, self-protection and protection of others were the most commonly reported reasons for mask use. In round 4, on the other hand, more than 50% of participants also reported wearing a mask due to requirements, either to enter a building or to receive a service (appendix A3). In rural areas, the proportion of mask users who reported wearing a mask to comply with a requirement nearly doubled between rounds 3 and 4. Fewer than 1% of respondents reported wearing a mask because they worried about fines imposed by the Government, because of social pressure or because they were sick or presented symptoms.

## DISCUSSION

In this study, we found that the magnitude of the SARS-CoV-2 outbreak in the country is likely larger than reflected in case-based surveillance datasets that rely on RT-PCR diagnostics. When we compared participants’ self-reports of a) symptoms often associated with COVID–19 (e.g., cough, chest pain) and b) their history of RT-PCR testing for SARS-CoV-2, we found that testing rates were low (approximately 5% of study participants). This was the case even among those who reported experiencing symptoms commonly associated with COVID–19 in clinical studies (e.g., cough, chest pain). As a result of very limited SARS-CoV-2 testing coverage, a number of cases might have remained undetected and were thus not included in daily tallies of cases compiled by the MoH.

This finding about the completeness of case counts derived from RT-PCR testing is affected by potential limitations. Our self-reported measure of prior SARS-CoV-2 testing, in particular, might misrepresent testing coverage. Due to stigma surrounding COVID–19 (86), some respondents might have failed to report previous tests for SARS-CoV-2. This would have prompted a downward bias in our estimates of testing coverage. Despite including a description of the sample collection procedure in our survey question in round 4, some participants might have confused SARS-CoV-2 testing with other prevention measures, e.g., temperature checks, screening questionnaires. This would have led to bias in the opposite direction, possibly inflating estimates of the proportion of participants tested for SARS-CoV-2. We also did not investigate repeat testing for SARS-CoV-2 among panel participants, nor did we assess whether testing coverage varied over time. Based on these data, and also owing to the selectivity of our study sample (see below), we could not produce an estimate of the true number of infections with SARS-CoV-2 in Malawi. Nonetheless, our findings corroborate results from a serosurvey conducted in Blantyre (87) and several similar surveys conducted in other African settings (12,88,89). In these studies, the high prevalence of IgG antibodies against SARS-CoV-2 implied a number of cases (much) larger than recorded by surveillance systems relying on RT-PCR testing.

Whereas our data indicated that daily case counts based on RT-PCR diagnostics might underestimate the scale of the outbreak, they also suggested that such laboratory-based surveillance systems might adequately capture epidemic trajectories. In particular, the panel data we collected on symptoms associated with COVID–19 indicated that an epidemic peak likely occurred in urban centers in June/July (figure 3), similar to trends detected in daily reports of PCR-confirmed cases (figure 1). At that time, the proportion of urban participants who reported symptoms commonly seen among COVID–19 cases (e.g., cough, chest pain) nearly doubled relative to levels observed in earlier months (i.e., round 1 in April/May). In subsequent rounds (i.e., rounds 3 and 4), this proportion rapidly returned to ‘”normal” levels. In contrast, the proportion of panel participants residing in rural areas and who reported a cough or another symptom, declined steadily from round to round, consistent with seasonal patterns of respiratory infections (90).

We found that several protective behaviors were adopted widely in our study sample during the first few months of the COVID-19 pandemic in Malawi. These included in particular behaviors that reduce the transmissibility of the novel coronavirus. More than half of panel participants reported adopting the recommended practice of physical distancing between rounds 1 & 3. Mask use also increased very sharply during that time frame, with more than 8 out of 10 urban participants reporting that they had worn a facial mask outside of their household in the past month (figure 8). Comparatively, behaviors that affect contacts between infected and susceptible individuals were more stable in our panel sample. Indicators of intra-household contact remained largely constant throughout the 4 panel rounds, and there were few changes in attendance of places and events where people socialize (and thus possibly spread SARS-CoV-2) during the course of the study

Our data on behavioral responses to the COVID–19 pandemic also indicate that panel participants have recently reduced their protective behaviors. For example, urban respondents are increasingly celebrating weddings (figure 6), a social occasion that had led to large SARS-CoV-2 clusters in other contexts (91). Most respondents have also reduced the consistency of their mask use: whereas in round 3, more than two thirds of mask users reported “always” wearing a mask outside of their household, this proportion dropped to <25% in round 4. Mask use appears increasingly sustained by requirements to wear masks to access various buildings and services, rather than by a motivation to protect others. These trends might enhance the likelihood of local transmission of SARS-CoV-2 in the event of new imported cases and/or the emergence of more transmissible viral strains (92).

These findings about the adoption of protective behaviors suffer from several limitations. First, respondents might have over-reported their use of recommended protective measures. For example, a recent study found that, in a sub-county of Kenya, self-reported mask use greatly exceeded the levels of mask use recorded by independent observers (93). In our study, the extent of such social desirability bias in self-reports of protective behaviors might have varied over time: incentives to report compliance with mask use recommendations might have been higher after the MoH had mandated the use of masks in public and imposed fines on non-compliers. Since this mandate was adopted when confirmed cases were near their peak (i.e., early August), this might have created a spurious temporal association between increasing reported mask use and declining case/symptoms. In our data however, the reported frequency of mask use declined sharply in round 4, at a time when the mandate to wear masks in public was still in place.

Second, our data on patterns of contact also lacked a proper baseline. Our first panel measurement took place in April/May, i.e., a few weeks after the Government declared COVID–19 a “national disaster”, closed schools and universities, and imposed limitations on the size of public gatherings. Reductions in attendance of social events (e.g., weddings) have likely occurred *before* our first data collection round. Such reductions in contact might have helped delay the start of the outbreak and reduce its magnitude. However, they did not fully prevent the local transmission of SARS-CoV-2: incidence increased sharply in June/July when restrictions on public gatherings were still in place. Our data on self-isolation are also limited because they was only collected in rounds 3 and 4. They indicated limited compliance with self-isolation guidelines, except among residents of urban areas who presented with a cough at the peak of the outbreak (figure 7). This factor might have contributed to slowing down the first wave of the pandemic in the country.

Third, there are several sampling limitations. Despite high enrollment and follow-up rates (figure 2), the sample size available for panel analyses was limited. We thus could not investigate the correlates of observed trends in symptoms and behaviors (e.g., gender differences, differences by occupation). Because it relied on mobile phones to conduct interviews, our sample was also selective. We thus reached more educated potential participants at a higher rate than potential participants with no schooling, or only primary education (78). If patterns of behavioral changes differed by educational levels, then our study results might be affected. Furthermore, the study sample was not representative of the population of Malawi, Karonga district or the urban areas of the country. It included members of families with ties to the area surrounding the lakeshore community of Chilumba and covered by the Karonga HDSS. These families were initially selected at random among household rosters compiled by the HDSS, during the pre-COVID–19 study. Their members are now dispersed throughout Malawi, including in the large cities most affected by the first wave of the COVID–19 pandemic in the country.

Finally, our analyses of behavioral changes can only suggest temporal associations with epidemic outcomes. In our data, reductions in the recorded incidence of SARS-CoV-2 (figure 1) and in self-reports of COVID–19 symptoms (figure 3) were preceded by, or occurred at approximately the same time as, the adoption of several protective behaviors (e.g., physical distancing, mask use). These apparent associations cannot be interpreted as causa, because we cannot rule out several competing explanations for this reversal of epidemic trends, due to data limitations. For example, the decline in incidence/symptoms of SARS-CoV-2/COVID–19 might have been prompted by successes in contact tracing, or changes in biological factors affecting the transmissibility of the novel coronavirus (e.g., temperature, humidity). Future investigations of the determinants of SARS-CoV-2 trends in Malawi and other African countries should triangulate multiple sources of data (e.g., behavioral interviews, viral sequences, serosurveys, contact tracing datasets).

Despite these limitations, our study adds to our understanding of the initial wave of the COVID–19 pandemic in Malawi. In highlighting that the number of persons infected during the first wave of the pandemic might be higher than recorded in surveillance systems that rely on RT-PCR testing, our work might help estimate the number of people who are still susceptible to the novel coronavirus more accurately. This might help strengthen projections of the impact of future waves of the COVID–19 pandemic in the country. Our results also emphasize the potential importance of population-level behavioral changes in containing SARS-CoV-2 outbreaks in Malawi. Highlighting the role that physical distancing and mask use might have played in containing the first COVID–19 wave might help sustain the implementation of these strategies in subsequent pandemic waves.

## Data Availability

At the moment, data are available upon reasonable request. The authors are preparing a data repository that will be made publicly available upon publication of study results.

**APPENDIX A1:**
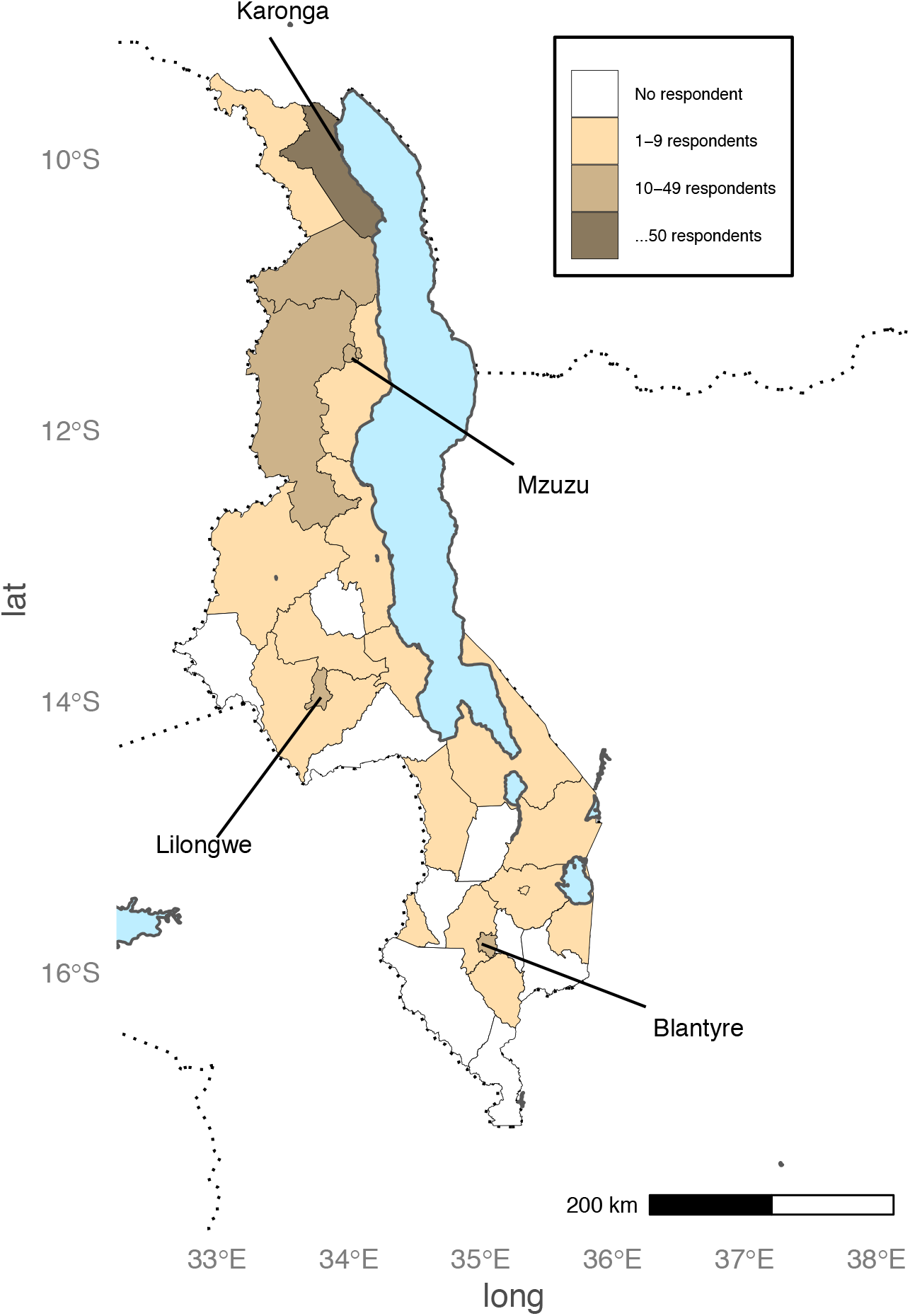
GEOGRAPHIC DISTRIBUTION OF RESPONDENTS. *Notes:* reported district of residence of study respondents during round 1. In our survey answer categories, Mzimba district was not split as Mzimba north vs. Mzimba south, as a result, we present data for these two districts jointly on the map. The city corporations of Lilongwe, Mzuzu, Zomba and Blantyre are displayed separately.

**APPENDIX A2:**
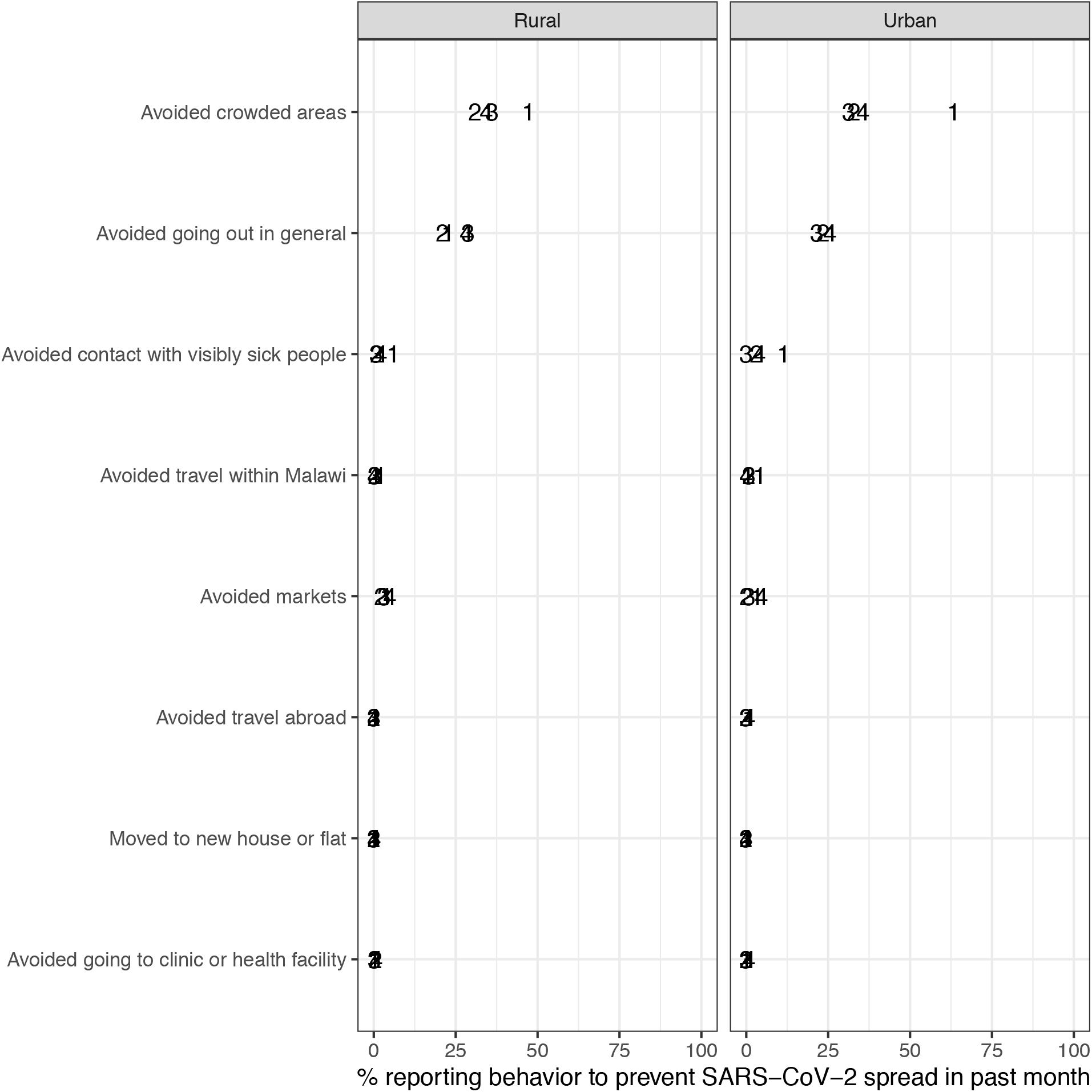
ADDITIONAL REPORTED BEHAVIORS TO LIMIT CONTACT BETWEEN INFECTED AND SUSCEPTIBLE INDIVIDUALS. *Notes:* the data plotted in this graph were collected from a question asking respondents what they had done in the past month to prevent the spread of SARS-CoV-2. Respondents were not provided with potential answers. However, after each response, interviewers were instructed to probe further, by asking respondents: “is there anything else that you have done?”. In this graph, the behaviors listed on the y-axis are those that reduce contact between infected and susceptible individuals. In each round, <1% of panel respondents reported not doing anything to prevent the spread of SARS-CoV-2. The point estimates are labeled by their round number on the plot. The behaviors that appear on the y-axis are listed according to their prevalence in urban areas in round 1.

**APPENDIX A3:**
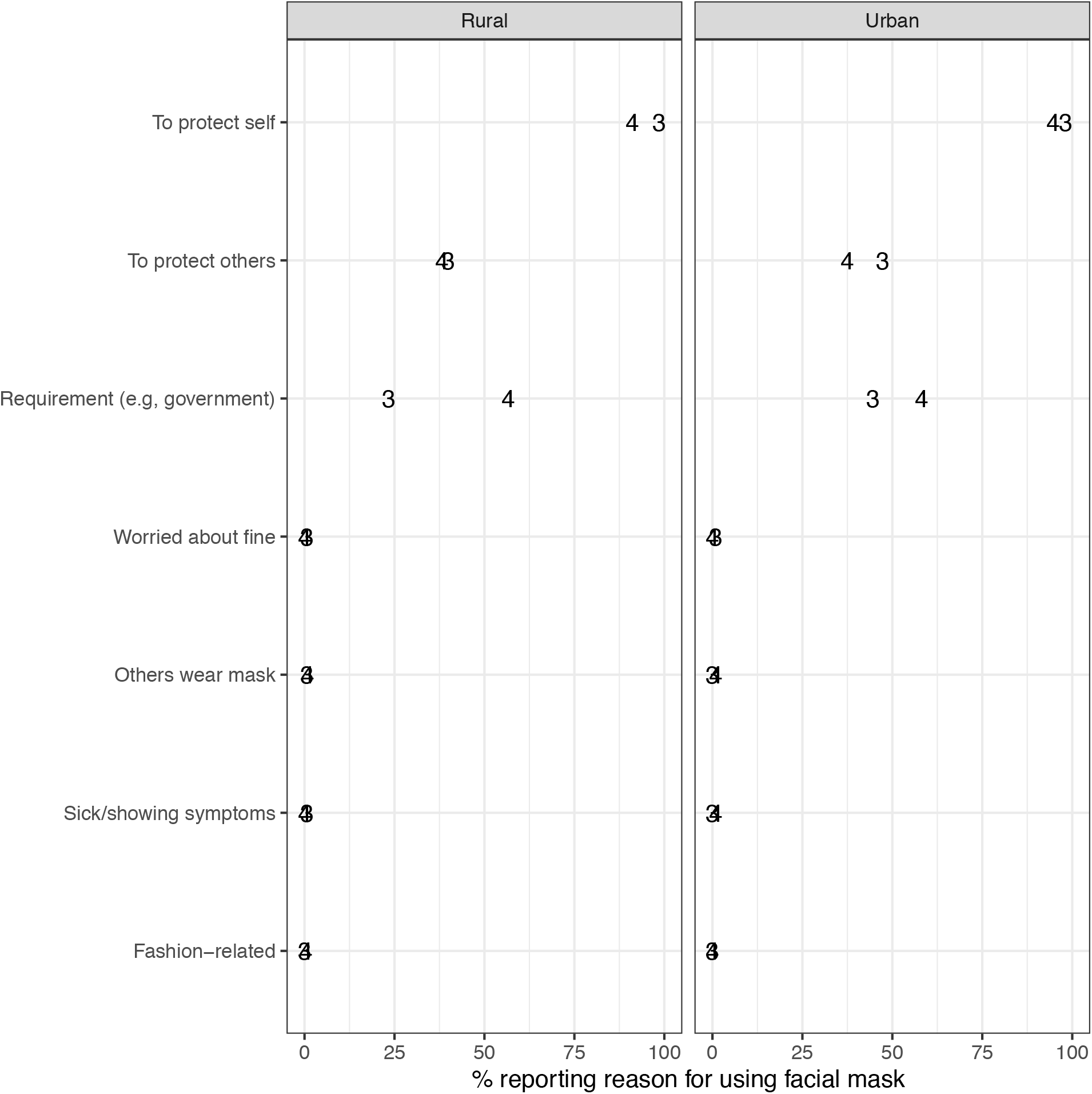
REPORTED REASONS FOR USING A MASK IN THE PAST MONTH. *Notes:* the data plotted in this graph were collected from a question asking respondents who reported having used a mask in the past month, why they had done so. Respondents were not provided with potential answers. However, after each response, interviewers were instructed to probe further, by asking respondents: “was there another reason?”. The point estimates are labeled by their round number on the plot. The reasons that appear on the y-axis are listed according to their prevalence in urban areas in round 1.

## Footnotes

6 Administrative capitals of each district

